# Loneliness in adults with cardiovascular disease and their social and emotional support needs: Implications for Hispanic adults from the 2023 Behavioral Risk Factor Surveillance System

**DOI:** 10.1101/2025.03.19.25324276

**Authors:** Derek S. Falk, Esmeralda Melgoza, Alberto Cabrera, Christian E. Vazquez

## Abstract

**Objectives:** Loneliness and social isolation pose significant risks for an individual’s physical, mental, and social health including higher incidence of cardiovascular disease (CVD), poorer patient reported outcomes, and earlier mortality compared to those not experiencing loneliness or social isolation. The objective of this study was to assess loneliness and social and emotional support among adults aged 18 years and older who have CVD in the US.

**Design:** Using the 2023 Behavioral Risk Factor Surveillance System’s social determinants and health equity module, we examined the distribution of US adults with CVD, compared the prevalence of CVD by Hispanic ethnicity, and conducted multivariable logistic regressions assessing the relationship of independent variables with loneliness and social and emotional support.

**Results:** The proportion of adults with CVD who felt lonely sometimes, usually, and always was 44.6%. Hispanic adults who felt lonely (56.3% vs. 43.0%; P<0.0001) and did not receive needed social and emotional support (13.7% vs. 9.8%; P=0.0162) experienced a higher prevalence of CVD than their non-Hispanic adult counterparts who felt lonely and did not receive needed social and emotional support. Adults with CVD who reported rarely or never receiving needed social and emotional support (odds ratio [OR]: 1.42; confidence interval [CI]: 1.14-1.77) had 42% higher odds of feeling lonely, compared to adults who indicated receiving social and emotional support sometimes, usually, or always. Among Hispanic adults with CVD, widowed/divorced/separated adults (OR: 2.30; CI: 1.46-3.61), urban residents (OR: 2.14; CI: 1.05-4.36), and unemployed adults (OR: 3.26; CI: 1.93-5.51) had higher odds of feeling lonely compared to married, rural, and employed adults.

**Conclusion:** This study demonstrates significant disparities in loneliness and social and emotional support in CVD among US adults, with Hispanics experiencing a disadvantage in both outcomes. Future studies should examine strategies to improve social connection for those experiencing disparities.

## Introduction

In 2023, the United States (U.S.) Surgeon General identified loneliness and social isolation as major public health problems (Office of the Surgeon General, 2023). Loneliness is the subjective internal state of distress that occurs from perceived isolation (Office of the Surgeon General, 2023). Social isolation is an objective measure of having few social relationships, social roles, group memberships, and infrequent social interactions (Office of the Surgeon General, 2023). In recent surveys, 58% of U.S. adults report experiencing loneliness (Buechler, 2021), 61% indicate that they do not have enough close friends or family, and 65% report feeling fundamentally disconnected from others (Making Caring Common, 2024). Loneliness and social isolation pose significant risks for an individual’s physical, mental, and social health (Office of the Surgeon General, 2023). Loneliness and social isolation are associated with lower self-esteem, increased risk of depression and anxiety, defective immune functioning, and higher blood pressure (Grant et al., 2009; Hawkley et al., 2010; Paul et al., 2021). Loneliness is also linked with higher incidence of cardiovascular (CVD) disease, poorer CVD patient outcomes, and earlier mortality from CVD compared to those who did experience loneliness (Paul et al., 2021).

CVD is the leading cause of morbidity and mortality in the US (Centers for Disease Control and Prevention, 2024c). CVD and its risk factors, however, are not evenly distributed across racial and ethnic groups, with marked disparities affecting socially disadvantaged and marginalized populations (Guadamuz et al., 2021). Hispanics, for example, have high rates of CVD-related risk factors, including obesity, hypertension, diabetes, and hyperlipidemia (Gomez et al., 2022). Hispanics also experience more systemic barriers linked to socioeconomic status (SES), including lower educational attainment, lower income, higher rates of unemployment, and worse living conditions compared to non-Hispanic Whites (Borkowski et al., 2024). High rates of food and housing insecurity, and a lack of adequate health insurance also contribute to worse CVD risk profiles among Hispanics compared to non-Hispanic Whites, which, in turn, contributes to high CVD prevalence in this population (Borkowski et al., 2024). Hispanic females and males have a total CVD prevalence of 42.7% and 52.3%, respectively (Gomez et al., 2022; Virani et al., 2021). Although Hispanics have a worse CVD risk profile, they have lower CVD-related mortality compared to non-Hispanic Whites, a phenomenon known as the Hispanic or Latino paradox (Balfour et al., 2016; Markides & Coreil, 1986).

Prior literature suggests that familism and collectivism partially explain the Hispanic or Latino paradox (Cahill et al., 2021; Lopez et al., 2023). Familism is defined as a key cultural value that emphasizes support, loyalty, honor, and obligation to the family (Cahill et al., 2021; Lopez et al., 2023). Increased levels of familism are associated with higher self-esteem, less loneliness, fewer depressive symptoms, fewer physical symptoms, more life satisfaction, and overall increased levels of well-being (Gallegos & Segrin, 2022). Several studies, however, also report that familism may also have a small but negative impact on an individual’s mental health, indicating that perhaps putting others’ welfare before the self may result in fear of burdening the family by disclosing health concerns or conditions and psychological distress (Corona et al., 2017; Gallegos & Segrin, 2022). Social and emotional support buffer some of the stressors that individuals may experience, and the opposite is true in the absence of social and emotional support. For example, a study reported that Latino immigrant men experience loneliness and low social support (Documet et al., 2019). Low social support among Latino immigrant men was associated with a higher frequency of binge drinking and depressive symptoms (Loury et al., 2011). Few studies have been able to assess loneliness among Hispanics in national datasets to contribute more evidence to the existence or absence of the Hispanic paradox in the context of CVD.

The objective of this study is to assess loneliness and social and emotional support among adults aged 18 years and older who have CVD in the US. We assess differences in loneliness and social and emotional support for the full-sample, and among Hispanics exclusively. This study advances the state of the literature by addressing the research gap that exists regarding loneliness and social and emotional support in the context of Hispanic adults with CVD. Our findings also inform future efforts to develop culturally tailored interventions.

## Methods

### Data source

The BRFSS is an annual, cross-sectional survey conducted by the Centers for Disease Control and Prevention (CDC) in coordination with all 50 states, the District of Columbia, and participating US territories to assess health risk behaviors, chronic conditions, and mental health using a random digit dialing approach for both landline and cell phones (Centers for Disease Control and Prevention, 2024b). Standard core questions are complemented by optional modules included in select states and territories. The 2023 social determinants and health equity module included responses from 39 states, the District of Columbia, and Puerto Rico. The present study examines responses from the social determinants of health module to address our research questions. Details of the design, sampling strategy, data collection, and weighting procedures have been provided by the CDC (Centers for Disease Control and Prevention, 2022).

## Measures

### Outcome variables

Two items from the social determinants and health equity module provided the outcome measures for our study. We measured the frequency of *loneliness* using the question “How often do you feel lonely?” and the frequency of *social and emotional support* with the question “How often do you get the social and emotional support you need?” Both items included a 5-point Likert scale with options including always, usually, sometimes, rarely, or never. We dichotomized the responses as always/usually/sometimes and rarely/never for both outcomes consistent with similar studies using these items (Bruss et al., 2024; Hacker et al., 2024).

### Cardiovascular disease

Our analyses only included a subset of adults with CVD. We used the recoded variable that assessed if respondents had ever reported coronary heart disease or myocardial infarction (yes/no) as an inclusion/exclusion variable for the analyses.

### Demographic variables

*Age* was categorized into 6 ranges from 18 to 24 years, 25 to 34 years, 35 to 44 years, 45 to 54 years, 55 to 64 years, and 65+ years. Race and ethnicity were two separate items that were subsequently recoded into a *race/ethnicity* variable with 5 categories: non-Hispanic Black, Hispanic, non-Hispanic multiracial, non-Hispanic other race, and non-Hispanic White. *Sex* included categories of male and female. *Marital status* was recoded into 3 groups consisting of widowed/divorced/separated, single, and married/member of an unmarried couple. *Language preference* was assessed based on the survey interview being in Spanish or English. The BRFSS provides an anonymized categorical variable denoting *residential location* as either rural or urban based on a recode of the respondents’ addresses. Respondents also indicated their *veteran status* as yes or no.

### Socioeconomic status

*Educational attainment* and *employment status* assessed socioeconomic status in our study. Educational attainment had the following categories: did not graduate high school, high school graduate, some college or technical school, and graduated from college or technical school. We recoded employment status into 3 categories that included: homemaker/student/retired, unemployed, and employed.

### Statistical analysis

We calculated the weights using all versions of the survey based on a previous description of analysis procedures for the BRFSS social determinants and health equity modules (Centers for Disease Control and Prevention, 2022). We analyzed the data using complex survey procedures incorporating these weights to estimate the corresponding population with CVD in locations included in the optional module. The univariable analysis described the frequency of the variables in the sample. Bivariable analyses assessed the prevalence of CVD by Hispanic ethnicity using Rao-Scott chi-square tests of categorical variables. Separate multivariable logistic regressions assessed loneliness and social and emotional support outcomes stratified by Hispanic ethnicity and adjusted by sociodemographic covariables. We conducted our analyses using SAS version 9.4 (SAS Institute, Inc., Cary, NC).

## Results

### Sample distribution

Table 1 shows the sample characteristics of U.S. adults with CVD who were 18 years and older. The incidence of CVD was highest among adults aged 65 years and older (56.7%), and lowest among adults aged 18 to 24 years (1.5%). Non-Hispanic White adults (66.7%), males (58.9%), individuals who were married or living as an unmarried couple (53.8%), lived in an urban setting (90.3%), and who were non-veterans (80.5%) were the majority in their respective categories. Educational attainment ranged from 17.2% of those who did not graduate high school to 30.7% who attended college. Employment status ranged from 21.4% of unemployed adults with CVD to 52.5% of those who were in the homemaker/student/retired category. Only 6.6% of respondents responded to the survey in Spanish compared to 93.4% in English. The proportion of adults with CVD who felt lonely (always/usually/sometimes) was 44.6%, while 10.3% rarely or never received the social and emotional support that they needed.

**Table 1.** Sample distribution of US adults aged 18+ years with cardiovascular disease and bivariable comparisons by Hispanic ethnicity.

|  | Full Sample<br>(N=25,698) |  | Non-Hispanic Adults<br>(n=24,110) |  | Hispanic Adults<br>(n=1,588) |  | P-Value |
| --- | --- | --- | --- | --- | --- | --- | --- |
|  | N | % | n | % | N | % |  |
| Age group |  |  |  |  |  |  | **<0.0001** |
| 18 to 24 years | 118 | 1.5 | 82 | 1.1 | 36 | 4.0 |  |
| 25 to 34 years | 302 | 2.6 | 223 | 2.2 | 79 | 6.0 |  |
| 35 to 44 years | 733 | 5.3 | 591 | 4.2 | 142 | 13.0 |  |
| 45 to 54 years | 1,879 | 11.3 | 1,601 | 10.4 | 278 | 17.6 |  |
| 55 to 64 years | 4,609 | 22.7 | 4,219 | 22.2 | 390 | 26.0 |  |
| 65+ years | 18,057 | 56.7 | 17,394 | 60.0 | 663 | 33.5 |  |
| Race/ethnicity |  |  |  |  |  |  | - |
| Non-Hispanic Black | 1,782 | 11.6 | 1,782 | 13.3 | - | - |  |
| Hispanic | 1,588 | 12.8 | - | - | 1,588 | 100.0 |  |
| Non-Hispanic, multiracial | 584 | 2.4 | 584 | 2.7 | - | - |  |
| Non-Hispanic, other race | 1,073 | 6.5 | 1,073 | 7.5 | - | - |  |
| Non-Hispanic White | 20,074 | 66.7 | 20,074 | 76.5 | - | - |  |
| Sex |  |  |  |  |  |  | 0.0842 |
| Male | 15,004 | 58.9 | 14,149 | 59.5 | 855 | 55.2 |  |
| Female | 10,694 | 41.1 | 9,961 | 40.5 | 733 | 44.8 |  |
| Marital status |  |  |  |  |  |  | **0.0005** |
| Widowed/divorced/separated | 10,349 | 35.5 | 9,746 | 35.2 | 603 | 37.9 |  |
| Single | 2,125 | 10.6 | 1,900 | 10.0 | 225 | 15.0 |  |
| Married/member of an unmarried couple | 13,051 | 53.8 | 12,300 | 54.8 | 751 | 47.1 |  |
| Survey language preference |  |  |  |  |  |  | **<0.0001** |
| Spanish | 869 | 6.6 | 32 | 0.1 | 837 | 51.8 |  |
| English | 24,829 | 93.4 | 24,078 | 99.9 | 751 | 48.2 |  |
| Residential location |  |  |  |  |  |  | **<0.0001** |
| Urban | 20,939 | 90.3 | 19,820 | 89.4 | 1,119 | 97.3 |  |
| Rural | 4,359 | 9.7 | 4,273 | 10.6 | 86 | 2.7 |  |
| Veteran status |  |  |  |  |  |  | **<0.0001** |
| Veteran | 5,831 | 19.5 | 5,663 | 21.1 | 168 | 8.3 |  |
| Non-veteran | 19,746 | 80.5 | 18,334 | 78.9 | 1,412 | 91.7 |  |
| Educational attainment |  |  |  |  |  |  | **<0.0001** |
| Did not graduate high school | 2,206 | 17.2 | 1,731 | 12.9 | 475 | 47.3 |  |
| Graduated high school | 7,378 | 29.9 | 6,952 | 30.8 | 426 | 22.9 |  |
| Some college or technical school | 7,502 | 30.7 | 7,141 | 32.2 | 361 | 20.3 |  |
| Graduated from college or technical school | 8,509 | 22.2 | 8,190 | 24.1 | 319 | 9.5 |  |
| Employment status |  |  |  |  |  |  | **<0.0001** |
| Homemaker/student/retired | 15,851 | 52.5 | 15,225 | 55.2 | 626 | 33.8 |  |
| Unemployed | 4,119 | 21.4 | 3,678 | 19.7 | 441 | 33.0 |  |
| Employed | 5,403 | 26.1 | 4,917 | 25.1 | 486 | 33.3 |  |
| Frequency of loneliness |  |  |  |  |  |  | **<0.0001** |
| Always/usually/sometimes | 10,400 | 44.6 | 9,583 | 43.0 | 817 | 56.3 |  |
| Rarely/never | 15,121 | 55.4 | 14,361 | 57.0 | 760 | 43.7 |  |
| Frequency of social and emotional support |  |  |  |  |  |  | **0.0162** |
| Rarely/never | 2,302 | 10.3 | 2,103 | 9.8 | 199 | 13.7 |  |
| Always/usually/sometimes | 22,981 | 89.7 | 21,606 | 90.2 | 1,375 | 86.3 |  |

Note: P values are Rao-Scott chi-square tests comparing the weighted proportions of non-Hispanic and Hispanic respondents for each variable and are bold where significant ( $P < 0.05$ ).

### Comparisons between non-Hispanic and Hispanic adults with CVD

The prevalence of CVD was 3 to 4 times higher for Hispanic adults versus non-Hispanic adults aged 18 to 24 years (4.0% vs. 1.1%), 25 to 34 years (6.0% vs. 2.2%), and 35 to 44 years (13.0% vs. 4.2%; P<0.0001). Hispanic adults also had higher prevalence of CVD for those aged 45 to 54 years (17.6% vs. 10.4%) and 55 to 64 years (26.0% vs. 22.2%) compared to non-Hispanic adults (P<0.0001). Hispanic single adults and those who were widowed/divorced/separated experienced higher prevalence of CVD compared to non-Hispanic adults who were single and widowed/divorced/separated (single: 15.0% vs. 10.0%; widowed/divorced/separated: 37.9% vs. 35.2%; P=0.0005). Survey language preference also varied significantly with 51.8% of Hispanic adults opting for the Spanish version compared to 0.1% of non-Hispanic adults (P<0.0001). Hispanic adults were predominantly urban residents at 97.3% compared to 89.4% of non-Hispanic adults (P<0.0001). Hispanic adults who did not graduate from high school had the highest prevalence of CVD at 47.3% compared to 12.9% of non-Hispanic adults who did not graduate from high school (P<0.0001).

Hispanic adults who were unemployed (33.0%) and employed (33.3%) had significantly higher prevalence of CVD than non-Hispanic adults who were unemployed (19.7%) and employed (25.1%; P<0.0001). Hispanic adults with CVD had a higher prevalence of feeling lonely (56.3%) compared to non-Hispanic adults with CVD (P<0.0001). Hispanic adults with CVD also had a higher prevalence of not receiving needed social and emotional support (13.7%) compared to non-Hispanic adults with inadequate social and emotional support (9.8%; P=0.0162).

### Multivariable logistic regressions of loneliness among US adults with CVD

We assessed the weighted and adjusted odds of feeling lonely (always/usually/sometimes) among US adults with CVD and performed a separate analysis for Hispanic adults with CVD (Table 2). Those aged 25 to 34 years (OR: 4.75; CI: 3.09-7.29) experienced the highest odds of feeling lonely compared to those aged 65+. Males with CVD, compared to females, experienced lower odds (OR: 0.87; 0.77-0.99) of reporting loneliness. Those who were widowed/divorced/separated (OR: 2.17; CI: 1.92-2.45), single (OR: 1.74; CI: 1.40-2.16), and urban residents (OR: 1.17; CI: 1.01-1.35) had greater odds of feeling lonely compared to married and rural residents.

**Table 2.** Odds of feeling lonely (always/usually/sometimes) among US adults with cardiovascular disease.

|  | All adults<br>(n=23,717) |  | Hispanic adults<br>(n=1,137) |  |
| --- | --- | --- | --- | --- |
|  | OR | 95% CI | OR | 95% CI |
| Age group |  |  |  |  |
| 18 to 24 years | **3.50** | **1.79-6.82** | **11.15** | **5.10-24.37** |
| 25 to 34 years | **4.75** | **3.09-7.29** | **7.28** | **2.32-22.82** |
| 35 to 44 years | **2.09** | **1.52-2.86** | **3.27** | **1.49-7.17** |
| 45 to 54 years | **1.63** | **1.30-2.06** | **2.98** | **1.59-5.58** |
| 55 to 65 years | **1.29** | **1.09-1.54** | **2.23** | **1.22-4.06** |
| 65+ years | Ref. |  | Ref. |  |
| Race/ethnicity |  |  |  |  |
| Non-Hispanic Black | 1.08 | 0.90-1.31 | - | - |
| Hispanic | 1.29 | 0.93-1.77 | - | - |
| Non-Hispanic, multiracial | 1.08 | 0.80-1.45 | - | - |
| Non-Hispanic, other race | **1.48** | **1.05-2.08** | - | - |
| Non-Hispanic White | Ref. |  | - | - |
| Sex |  |  |  |  |
| Male | **0.87** | **0.77-0.99** | 1.00 | 0.67-1.49 |
| Female | Ref. |  | Ref. |  |
| Marital status |  |  |  |  |
| Widowed/divorced/separated | **2.17** | **1.92-2.45** | **2.30** | **1.46-3.61** |
| Single | **1.74** | **1.40-2.16** | 1.37 | 0.83-2.28 |
| Married/member of an unmarried couple | Ref. |  | Ref. |  |
| Survey language preference |  |  |  |  |
| Spanish | 0.97 | 0.61-1.54 | 1.05 | 0.63-1.74 |
| English | Ref. |  | Ref. |  |
| Residential location |  |  |  |  |
| Urban | **1.17** | **1.01-1.35** | **2.14** | **1.05-4.36** |
| Rural | Ref. |  | Ref. |  |
| Veteran status |  |  |  |  |
| Veteran | 1.08 | 0.93-1.25 | 1.17 | 0.58-2.36 |
| Non-veteran | Ref. |  | Ref. |  |
| Educational attainment |  |  |  |  |
| Did not graduate high school | **1.35** | **1.10-1.67** | 1.01 | 0.56-1.84 |
| Graduated high school | 1.09 | 0.94-1.26 | 0.63 | 0.35-1.11 |
| Some college or technical school | 1.05 | 0.90-1.21 | 0.88 | 0.49-1.60 |
| Graduated from college or technical school | Ref. |  | Ref. |  |
| Employment status |  |  |  |  |
| Homemaker/student/retired | **1.30** | **1.08-1.55** | **3.81** | **2.08-6.96** |
| Unemployed | **2.25** | **1.86-2.71** | **3.26** | **1.93-5.51** |
| Employed | Ref. |  | Ref. |  |
| Frequency of social and emotional support |  |  |  |  |
| Rarely/never | **1.42** | **1.14-1.77** | 1.12 | 0.58-2.16 |
| Always/usually/sometimes | Ref. |  | Ref. |  |

Note: OR=Odds Ratio; CI=Confidence Interval. Significant results (95% CI does not contain 0) are bold for emphasis.

Respondents with CVD who did not graduate high school (OR: 1.35; CI: 1.10-1.67), identified as homemaker/student/retired (OR: 1.30; CI: 1.08-1.55), and unemployed (OR: 2.25; CI: 1.86-2.71) also had higher odds of feeling lonely compared to adults with CVD who graduated from college and were employed. Adults who rarely or never received social and emotional support had 42% higher odds of loneliness (OR: 1.42; CI: 1.14-1.77) compared to adults who sometimes, usually or always received needed social and emotional support.

Among Hispanic adults with CVD, the odds of feeling lonely were even higher compared to the full sample. Individuals aged 18 to 24 years (OR: 11.15; CI: 5.10-24.37) had the highest odds of feeling lonely, while individuals aged 55 to 64 (OR: 2.23; CI: 1.22-4.06) years had the lowest odds compared to those aged 65+ (Table 2).

Widowed/divorced/separated adults (OR: 2.30; CI: 1.46-3.61), urban residents (OR: 2.14; CI: 1.05-4.36), homemaker/student/retired adults (OR: 3.81; CI: 2.08-6.96), and unemployed adults (OR: 3.26; CI: 1.93-5.51) had higher odds of feeling lonely compared to adults who were married/member of an unmarried couple, rural residents, and employed.

### Multivariable logistic regressions of social and emotional support among US adults with CVD

We assessed the odds of not receiving needed social and emotional support (rarely/never) among the full US adult sample with CVD and separately for Hispanic adults (Table 3). Males with CVD (OR: 1.44; CI: 1.15-1.80) had higher odds of not receiving needed social and emotional support compared to females. Those aged 25 to 34 years (OR: 2.41; CI: 1.35-4.30), 35 to 44 years (OR: 1.69; CI: 1.08-2.66), 45 to 54 years (OR: 1.46; CI: 1.02-2.09), non-Hispanic multiracial adults (OR: 2.20, CI: 1.41-3.45), non-Hispanic adults of another race (OR: 2.22; CI: 1.41-3.49), widowed/divorced/separated (OR: 1.78; CI: 1.42-2.23), and single (OR: 1.80; CI: 1.30-2.49) had higher odds of not receiving needed social and emotional support compared to those with CVD who were 65+, non-Hispanic White, married/living as an unmarried couple. Respondents who did not graduate high school (OR: 1.72; CI: 1.16-2.53) and high school graduates (OR: 1.44, CI: 1.08-1.90) had higher odds of not receiving needed social and emotional support compared to college graduates with CVD. Unemployed adults with CVD (OR: 1.63; CI: 1.24-2.14) had 63% higher odds of not receiving needed social and emotional support compared to employed adults with CVD.

**Table 3.** Odds of not receiving needed social and emotional support (rarely/never) among adults in the US with cardiovascular disease.

|  | All adults<br>(n=23,831) |  | Hispanic adults<br>(n=1,144) |  |
| --- | --- | --- | --- | --- |
|  | OR | 95% CI | OR | 95% CI |
| Age group |  |  |  |  |
| 18 to 24 years | 1.71 | 0.64-4.57 | 2.32 | 0.75-7.19 |
| 25 to 34 years | **2.41** | **1.35-4.30** | 1.46 | 0.29-7.26 |
| 35 to 44 years | **1.69** | **1.08-2.66** | 2.37 | 0.64-8.78 |
| 45 to 54 years | **1.46** | **1.02-2.09** | 1.51 | 0.47-4.81 |
| 55 to 64 years | 1.25 | 0.90-1.73 | 2.00 | 0.61-6.56 |
| 65+ years | Ref. |  | Ref. |  |
| Race/ethnicity |  |  |  |  |
| Non-Hispanic Black | 1.11 | 0.83-1.48 | - | - |
| Hispanic | 1.35 | 0.78-2.34 | - | - |
| Non-Hispanic, multiracial | **2.20** | **1.41-3.45** | - | - |
| Non-Hispanic, other race | **2.22** | **1.41-3.49** | - | - |
| Non-Hispanic White | Ref. |  | - | - |
| Sex |  |  |  |  |
| Male | **1.44** | **1.15-1.80** | **2.25** | **1.26-4.03** |
| Female | Ref. |  | Ref. |  |
| Marital status |  |  |  |  |
| Widowed/divorced/separated | **1.78** | **1.42-2.23** | 0.73 | 0.39-1.35 |
| Single | **1.80** | **1.30-2.49** | 0.86 | 0.37-1.99 |
| Married/member of an unmarried couple | Ref. |  | Ref. |  |
| Survey language preference |  |  |  |  |
| Spanish | 0.95 | 0.44-2.05 | 1.05 | 0.50-2.17 |
| English | Ref. |  | Ref. |  |
| Residential location |  |  |  |  |
| Urban | 0.94 | 0.75-1.19 | 2.04 | 0.95-4.38 |
| Rural | Ref. |  | Ref. |  |
| Veteran status |  |  |  |  |
| Veteran | 1.03 | 0.79-1.35 | 1.75 | 0.83-3.72 |
| Non-veteran | Ref. |  | Ref. |  |
| Educational attainment |  |  |  |  |
| Did not graduate high school | **1.72** | **1.16-2.53** | **4.36** | **1.66-11.48** |
| Graduated high school | **1.44** | **1.08-1.90** | **3.91** | **1.63-9.38** |
| Some college or technical school | 1.18 | 0.90-1.56 | 2.13 | 0.89-5.12 |
| Graduated from college or technical school | Ref. |  | Ref. |  |
| Employment status |  |  |  |  |
| Homemaker/student/retired | 1.31 | 0.95-1.80 | 1.61 | 0.58-4.45 |
| Unemployed | **1.63** | **1.24-2.14** | 0.77 | 0.38-1.57 |
| Employed | Ref. |  | Ref. |  |

Note: OR=Odds Ratio; CI=Confidence Interval. Significant results (95% CI does not contain 0) are bold for emphasis.

In the separate analysis for Hispanic adults only, males with CVD (OR: 2.25; CI: 1.26-4.03) had over 2 times the odds of not receiving needed social and emotional support compared to females. Those who did not graduate high school (OR: 4.36; CI: 1.66-11.48) and high school graduates (OR: 3.91; CI: 1.63-9.38) also had increased odds of not receiving needed social and emotional support compared to college graduates with CVD.

## Discussion

This study assessed loneliness and social and emotional support outcomes among adults who have CVD in the US, with an emphasis on differences between Hispanics and non-Hispanics. The findings suggest Hispanics with CVD have higher rates of feeling lonely and not receiving the support they need, compared to non-Hispanics. Hispanic adults had higher prevalence of CVD than non-Hispanic adults. Hispanic adults, compared to non-Hispanic adults, who felt lonely and did not receive needed social and emotional support also experienced a higher prevalence of CVD. There were more statistically significant findings in the full sample analyses compared to the Hispanic sample only for both outcomes; however, the effect size for any matching finding was always higher for the Hispanic sample compared to the full sample. Overall, the findings align with literature that Hispanics have higher at-risk profiles for all key variables, especially at the intersection of loneliness, social and emotional support, and CVD.

The literature highlights the interconnection between social support and loneliness, such that social support may partially buffer against the negative impacts of loneliness for Hispanics via cultural values such as collectivism and familism (Lee et al., 2020). However, the current sample shows low rates of both outcomes which can be linked to the CVD context. Though the literature has mixed findings related to rates of loneliness and social support for Hispanics compared to non-Hispanics, most studies suggest that when loneliness does occur in Hispanic adults, it is associated with elevated risks of metabolic and cardiovascular disorders compared to other groups (Foti et al., 2020; Tibirica et al., 2022). Another study using the 2022 BRFSS identified disparities in social isolation and social support particularly for Hispanic adults with lower educational attainment and higher unemployment rates, which is consistent with the current study (Bruss et al., 2024). It is possible that the potential protective factors against loneliness and lack of social support may not be in place for those with CVD, leading to higher likelihood of loneliness and lack of social support.

There are several studies that also found younger adults with CVD are at high-risk of being lonely due to the life stage of still building a network, friendships, and other relationships (Carter et al., 2015; Christiansen et al., 2021; McGlone & Long, 2020).

Having a chronic disease that interrupts this period marked by social activities may exacerbate loneliness. There is also a stigma of being undesirable as a friend or partner with CVD (Carter et al., 2015; McGlone & Long, 2020), where older age groups may have learned to cope or have established networks and relationships by later life. For Hispanic younger adults, the layering of identity can make this more challenging. For example, the anti-immigrant environment has increased in recent years, creating generalized fear and alienation, even if not an immigrant but may be perceived to be, that may prevent Hispanics from attaining the supports they need (Garcia et al., 2022; Lee et al., 2020). They may feel isolated from Hispanic older networks, especially if they do not speak Spanish, leaving them to straddle both cultures and not quite fitting in with a group for support.

Our findings confirm other studies that have found that urban residents experience higher rates of loneliness than rural residents (Pickering et al., 2023). Several studies have demonstrated that older adults in rural settings are more engaged in social activities in their daily lives and were partially protected during the COVID-19 pandemic by adapting more quickly to digital technologies that reduced social isolation, being able to perform activities such as gardening that distanced them from others, and engaging in volunteer activities (Pickering et al., 2023). Rural disadvantages were associated with living alone, being a caretaker, and having chronic health conditions or substance misuse problems (Pickering et al., 2023). Others have found that rural residents enjoy more social relationships and are able to rely on family connections compared to urban residents, although racial and ethnic minorities in rural areas did not experience the same benefits (Henning-Smith et al., 2019).

The current findings also align with the literature indicating that Hispanic men experience low social support (Garcia et al., 2022). This is partly due to barriers that are pronounced for Hispanic men, who face expectations of traditional masculine norms that may discourage help-seeking behaviors. The pattern of Hispanic men being reticent to engage in behaviors such as sharing emotions, reaching out for help, or engaging in interventions has been documented for nearly two decades (Gonzalez et al., 2022; Shattell et al., 2008). Anecdotally, the authors of the current study engage in conversations with other researchers across the US to continue to learn best practices to engage Hispanic men for research and practice, and this is shared to highlight that this is an ongoing challenge nationally.

### Strengths and Limitations

Our study has several strengths including the use of a survey that includes data on health-related risk behaviors, chronic health conditions, and use of preventive services across 39 states, the District of Columbia, and Puerto Rico. This study is also not without limitations. The first limitation is the BRFSS response rate. In 2023, the BRFSS response rate was 44.7%, which although low, is comparable with other surveys in the U.S. (Centers for Disease Control and Prevention, 2024a). For example, response rates for the California Health Interview Survey and the National Health Interview Survey were 11.2% and 61.1%, respectively. The second limitation is the reliance of BRFSS on respondent self-report which may introduce recall bias. A third limitation is the cross-sectional design of this study, which limits our ability to establish causality. A fourth limitation is the exclusion of certain populations from the BFRSS variables. These populations are often the most marginalized and have a higher prevalence of loneliness and social isolation. For example, sex at birth is defined as male and female, excluding non-binary populations due to small sample sizes particularly in the analyses considering Hispanics only. A previous study, however, reported that loneliness and social isolation were the highest among transgender populations (Bruss et al., 2024).

## Conclusion

This study demonstrates significant disparities in loneliness and social and emotional support in CVD among US adults, with Hispanics experiencing a disadvantage in both outcomes. The findings highlight the cumulative risk for younger, unemployed individuals with lower educational attainment, particularly among Hispanics. Culturally and linguistically tailored interventions are needed to support at risk Hispanic adults with CVD to mitigate the impact of loneliness and lack of social and emotional support for this group. Future studies should examine strategies to improve social connection through access to supportive networks while addressing barriers contributing to these disparities. These efforts may help improve health, mental health, and social outcomes for those with CVD, particularly for Hispanic adults.

## Data Availability

The data that support the findings of this study are openly available at https://www.cdc.gov/brfss/.

## Ethical approval statement

Ethical approval was not required for this study as all data are deidentified, publicly available and deemed not human subjects research.

